# Clinically Applicable Deep Learning for Intracranial Aneurysm Detection in Computed Tomography Angiography Images: A Comprehensive Multicohort Study

**DOI:** 10.1101/2020.03.21.20040063

**Authors:** Zhao Shi, Chongchang Miao, Chengwei Pan, Xue Chai, Xiu Li Li, Shuang Xia, Yan Gu, Yonggang Zhang, Bin Hu, Wenda Xu, Changsheng Zhou, Son Luo, Hao Wang, Li Mao, Kongming Liang, Yizhou Yu, Guang Ming Lu, Long Jiang Zhang

**Affiliations:** Department of Medical Imaging, Jinling Hospital, Medical School of Nanjing University, Nanjing, Jiangsu, 210002, P.R. China; Department of Radiology, Lianyungang First People’s Hospital, Lianyungang, Jiangsu, 222002, P.R. China; Computer Science Department, School of EECS, Peking University, Beijing, 100089, P.R. China; Department of Radiology, Nanjing Brain Hospital, Nanjing, Jiangsu, 210002, P.R. China; DeepWise AI lab. Beijing, 100089, P.R. China; Department of Radiology, Tianjin First Central Hospital, Tianjin, 300192, P.R. China; Department of Medical Imaging, Jinling Hospital, Xuzhou Medical University, Xuzhou, Jiangsu, 221004, P.R. China

## Abstract

Intracranial aneurysm is a common life-threatening disease. CTA is recommended as a standard diagnosis tool, while the interpretation is time-consuming and challenging. We presented a novel deep-learning-based framework trained on 1,177 DSA verified bone-removal CTA cases. The framework had excellent tolerance to the influence of occult cases of CTA-negative but DSA-positive aneurysms, image quality, and manufacturers. Simulated real-world studies were conducted in consecutive internal and external cohorts, achieving improved sensitivity and negative predictive value than radiologists. A specific cohort of suspected acute ischemic stroke was employed and found 96.8% predicted-negative cases can be trusted with high confidence, leading to reducing in human burden. A prospective study is warranted to determine whether the algorithm could improve patients’ care in comparison to radiologists’ assessment.

## Introduction

Intracranial aneurysms (IAs) are relatively common life-threatening diseases with a prevalence of 3.2% in general population^1^ and account for 80-85% in spontaneous subarachnoid hemorrhage (SAH) population^2^. IAs are increasingly being detected due to the widespread application of advanced imaging techniques. Although IAs related SAH accounts for 5-10% of all strokes in the United States^3^, it may cause significantly high mortality^4^, and the survivors may suffer from long-term neuropsychological effects and decreased quality of life^5^. Early diagnoses of underlying IA can both influence clinical management and guide prognosis in intracerebral hemorrhage patients^7^. For patients with spontaneous SAH, timely and accurate identification of IAs is critical for immediate interventional or surgery management^8^, while for patients without IAs, reliable exclusion of IAs is also important for specialized management.

Computed tomographic angiography (CTA) is a noninvasive, convenient, and reliable modality to detect IAs^9^. AHA/ASA guidelines have recommended CTA as a useful tool for detection and follow-up of unruptured IAs (Class I; Level B)^10^ and the workup of aneurysmal SAH (Class IIb, Level C)^11^. CTA is the standard-of-care imaging modality for the patients suspicious of SAH from the rupture of IAs in emergency department. However, CTA interpretation is time-consuming and subspecialty-training-requiring. The existing challenges also include inter-grader variability and high false-positive (FP), false-negative (FN) rates. The diagnostic accuracy is dependent on several factors including aneurysms size, diversity of technological specifications (16-versus 64-detector rows), image acquisition protocols, image quality, image postprocessing algorithms and variations in radiologists’ experiences, resulting in a mean sensitivity in the range of 70.7%–97.8% in detecting IAs^9,12^. The recently published guideline of acute ischemic stroke (AIS) has strongly recommended CTA (with CT perfusion) use (Class I and level A) for selecting candidates for mechanical thrombectomy between 6 and 24 hours after illness onset ^13-15^. Consequently, the workload of the radiologists to detect or exclude IAs is rapidly increasing in the non-SAH setting, where excluding IAs on CTAs remains a challenging task^16^. Given all the preconditions mentioned above, it is timely and urgently needed to have high-performance computer assisted diagnosis (CAD) tools to help detect, increase efficiency and reduce disagreement among observers, finally potentially improving clinical care of the patients.

MR angiography (MRA) or CTA based CAD programs have been devised to automatically detect IAs. The conventional-style CAD systems were based on pre-supplied characteristics or imaging features, such as vessel curvature, thresholding, or a region-growing algorithm^17,18^, while the performance and generalization are not satisfactory. Nowadays, deep learning (DL) has shown significant potential in accurately detecting lesions on medical imaging and had reached or even superior to the expertise-level of diagnosis^19-22^. DL is a machine learning technique that directly learns the most predictive features from a large data set of labeled images^21,22^. Explorations on DL combined with MRA have reported decent results for IA detection^23,24^. While CTA based CAD system has been rarely reported, and only two recently published studies can be found, to the best of our knowledge^25,26^. Notably, these models were trained in small sample size, without the gold standard, and were not tested in different scenarios, thus they were not adequate to apply in real-world clinical settings, which may fall into the ‘AI chasm’^27^. It is necessary and imperative to develop a robust and reliable AI tool for CAD of IAs in a clinical real-world application. The experiments presented in this article will endeavor to resolve these problems.

To properly address the shortcomings of current computational approaches in the context of IAs detection and enable the clinical deployment, it requires training and validation of models on large-scale datasets representative of the wide variability of cases encountered in the routine clinic. Therefore, we collected 1,177 head bone-removal CTA images with/without SAH to derive a newly proposed framework for automated detection of IAs, in a way we assume it can simulate the detecting procedure of the human brain. Furthermore, we aimed to ascertain the capability and generalizability of the framework using real-world CTA images from 4 hospitals and to test the framework in a simulated real-world routine clinical setting. In order to clearly identify the presence of IAs, especially small ones, we only enrolled the patients who had underwent cerebral digital subtraction angiography (DSA), the gold standard for diagnosing IAs, to verify the results of CTA in the training dataset.

The novel framework was an end-to-end 3D CNN segmentation model (**Fig. 1**). First, an encoder-decoder architecture was used for smooth and gradual transitions from original images to segmentation mask. Secondly, residual blocks^28^ were adopted to make the stable training for increasing depth of the network. Third, a dual-attention^29^ block was embedded to learn long-range contextual information to get more reliable feature representations. Besides one cohort for model training, we also made a comprehensive analysis of the proposed framework on the effects of different factors and in real-world clinical scenarios by 7 other cohorts (**Table 1**).

**Table 1.** Overview of the baseline characteristics of the 8 cohorts

|  | Internal cohort<br>1, n=1177 | Internal cohort<br>2, n=245 | Internal cohort<br>3, n=151 | Internal cohort<br>4, n=374 | Internal cohort<br>5, n=214 | NBH cohort,<br>n=211 | TJ cohort,<br>n=59 | LYG cohort,<br>n=316 |
| --- | --- | --- | --- | --- | --- | --- | --- | --- |
| **Patients with IAs, n (%)** | 869 (73.8) | 111 (45.3) | 46 (30.5) | 53 (14.2) | 10 (4.7) | 39 (18.5) | 39 (66.1) | 60 (19.0) |
| **Number of IAs, n** | 1099 | 148 | 59 | 71 | 12 | 47 | 50 | 76 |
| **Patients without IAs, n (%)** | 308 (26.2) | 134 (54.7) | 105 (69.5) | 321 (85.8) | 204 (95.3) | 172 (81.5) | 20 (33.9) | 256 (81.0) |
| **Male sex, n (%)** | 311 (28.3) | 136 (55.5) | 84 (55.6) | 242 (64.7) | 150 (70.1) | 145 (68.7) | 32 (54.2) | 187 (59.2) |
| **SAH, n (%)** | 931 (79.1) | 108 (44.1) | 59 (39.1) | 28 (7.5) | 0 (0) | 5 (2.4%) | 32 (54.2) | 47 (14.9) |
| **Number of patients with IAs, n (%)** | 760 (81.6) | 87 (80.6) | 39 (66.1) | 10 (35.7) | 0 (0) | 2 (40.0%) | 30 (93.8) | 25 (53.2) |
| **Non SAH (IA %)** | 246 (20.9) | 137 (54.9) | 92 (60.9) | 346 (92.5) | 214 (29.9) | 206 (97.6%) | 27 (45.8) | 269 (85.1) |
| **Number of patients with IAs, n (%)** | 109 (44.3) | 24 (17.5) | 7 (7.6) | 43 (12.4) | 10 (4.9) | 37 (18.0%) | 9 (33.3) | 35 (13.0) |
| **age, years** | 54.0<br>(46.0,62.0) | 59.0<br>(48.0,66.0) | 60.0<br>(51.5,69.0) | 63.0<br>(50.0,70.0) | 68.2±19.5 | 60.2±9.9 | 58.5±11.6 | 65.5<br>(52.5,73.0) |
| **age<70 years, n (%)** | 1088 (92.4) | 209 (85.3) | 117 (77.5) | 273 (73.0) | 130 (60.7) | 150 (71.1) | 49 (83.1) | 223 (70.6) |
| **Location** |  |  |  |  |  |  |  |  |
| **MCA, n (%)** | 131 (11.9) | 25 (16.9) | 6 (10.2) | 13 (18.3) | 3 (25.0) | 3 (6.4) | 11 (22.0) | 13 (17.1) |
| **ACoA, n (%)** | 291 (26.5) | 29 (19.6) | 14 (23.7) | 8 (11.3) | 0 (0) | 5 (10.6) | 8 (16.0) | 10 (13.2) |
| ICA, n (%) | 207 (18.8) | 34 (23.0) | 12 (20.3) | 15 (21.1) | 7 (58.3) | 22 (46.8) | 6 (12.0) | 19 (25.0) |
| PCoA, n (%) | 322 (29.3) | 33 (22.3) | 21 (35.6) | 19 (26.8) | 2 (16.7) | 5 (10.6) | 13 (26.0) | 19 (25.0) |
| VBA, n (%) | 40 (3.6) | 7 (4.7) | 0 (0) | 9 (12.7) | 0 (0) | 8 (17.0) | 9 (18.0) | 7 (9.2) |
| CA, n (%) | 43 (3.9) | 6 (4.1) | 2 (3.4) | 3 (4.2) | 0 (0) | 0 (0) | 1 (2.0) | 1 (1.3) |
| ACA, n (%) | 41 (3.7) | 11 (7.4) | 3 (5.1) | 4 (5.6) | 0 (0) | 3 (6.4) | 0 (0) | 5 (6.6) |
| PCA, n (%) | 24 (2.3) | 3 (2.0) | 1 (1.7) | 0 (0) | 0 (0) | 1 (2.2) | 2 (4.0) | 2 (2.6) |
| **Size** |  |  |  |  |  |  |  |  |
| <3, n (%) | 204 (18.6) | 32 (21.6) | 17 (28.8) | 22 (31.0) | 1 (8.3) | 9 (19.1) | 6 (12.0) | 18 (23.7) |
| ≥3, <5, n (%) | 426 (38.8) | 46 (31.1) | 23 (39.0) | 25 (35.2) | 3 (25.0) | 17 (36.2) | 19 (38.0) | 30 (39.5) |
| ≥5, <10, n (%) | 401 (36.5) | 65 (43.9) | 15 (25.4) | 16 (22.5) | 6 (50.0) | 18 (38.3) | 20 (40.0) | 21 (27.6) |
| ≥10, n (%) | 68 (6.1) | 5 (3.4) | 4 (6.8) | 8 (11.3) | 2 (16.7) | 3 (6.4) | 5 (10.0) | 7 (9.2) |
| size, mm | 4.3 (3.0,6.0) | 4.8 (3.3,6.3) | 4.3 (2.8,5.2) | 3.5 (2.9,6.5) | 5.8±2.8 | 4.3 (3.3,6.7) | 5.0 (4.0,6.0) | 4.4 (3.2,6.2) |

*ACA, anterior cerebral artery; ACoA, anterior communication artery; CA, cerebellar artery; IA, intracranial aneurysm; ICA, internal carotid artery; MCA, middle cerebral artery; PCA, posterior cerebral artery; PCoA, posterior communication artery; SAH, subarachnoid hemorrhage; VBA, vertebral basilar artery.*

**Fig.1.**
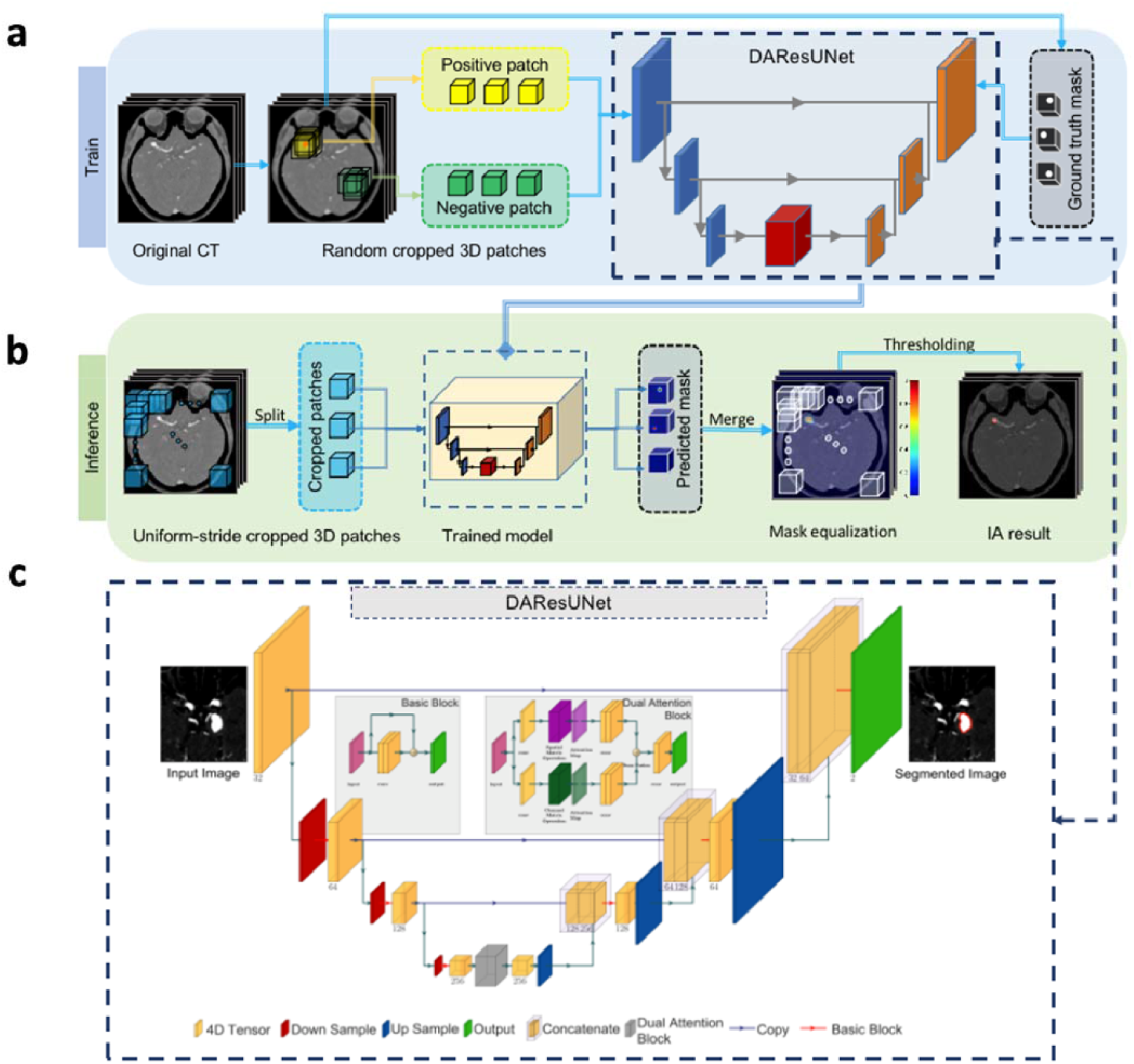
Overview of the proposed DL framework presented in this study. **a**, Training stage: we used 3D patches randomly sampled from the bone-removal CTA scans to train the network. **b**, Inference stage: Uniform-stride sampling was used and then the prediction of those samples was merged to get the final prediction of the whole volume. **c**, Illustration of the architecture of the end-to-end aneurysm prediction framework. The proposed segmentation model had a similar encoder-decoder architecture as U-Net^30^, and residual blocks and a dual attention block were used to improve the performance of the network.

## Results

### Model Development and Primary Validation

There were 16,975 consecutive head or head/neck CTA and 7,035 cerebral DSA performed between Jun. 1, 2009 and Mar. 31, 2017 in Jinling Hospital, among which 1,875 patients underwent both CTA and DSA. After quality control evaluation and images review, the final dataset (**Internal cohort 1**) consisted of **371**,**932** images from **1**,**177** cases (316 slices/case): 869 patients with 1099 aneurysms and 308 non-aneurysm controls, including 257 patients without abnormal findings and 51 patients with intracranial artery stenosis. Approximately 5 representative slices out of the 316 slices of the scan were manually segmented (2-47 slices). The cohort was split into training/tunning/testing sets. The training set included 927 cases (744 cases with IAs and 183 non-aneurysm controls); the tunning set consisted 100 cases (50 cases with IAs and 50 controls); the testing set had 150 cases with 50% cases having IAs **(Extended Data Fig. 1)**.

The DL algorithm was developed on the training set. After the entire training procedure (see **Methods**), the model with the best recall rate on the tunning set was cherry-picked. Furthermore, the sensitivity and specificity of the framework under different FPs were analyzed **(Extended Data Fig. 2)**, and the comprehensive optimal threshold was determined to make the FPs to be 0.3 so that the framework reached a high sensitivity of 97.3% with a moderate specificity of 74.7% on the testing set. For aneurysm-based analysis, the recall rate was 95.6% with a Dice index of 0.752. Totally 4 aneurysms from 4 patients were missed, including 2 patients with multiple IAs. All of the missed aneurysms were small (< 5 mm), among which 3 were tiny IAs (≤ 3 mm), 2 were located in the cerebellar artery (CA). That is the framework reaching 100% recall rate for aneurysms located in anterior cerebral artery (ACA), middle cerebral artery (MCA), anterior communication artery (ACoA), vertebral basilar artery (VBA) and posterior cerebral artery (PCA), and 99.5% for aneurysms located in internal carotid artery (ICA) and posterior communication artery (PCoA), 95.3 % for CA; 100% recall rate for aneurysms ≥ 5 mm, 99.9% for those ≥ 3 mm, 99.6% for all aneurysms. The framework took a mean of 17.6 seconds (95% CI: 17.2-18.0 seconds) to process an examination and output its segmentation map.

**Fig.2.**
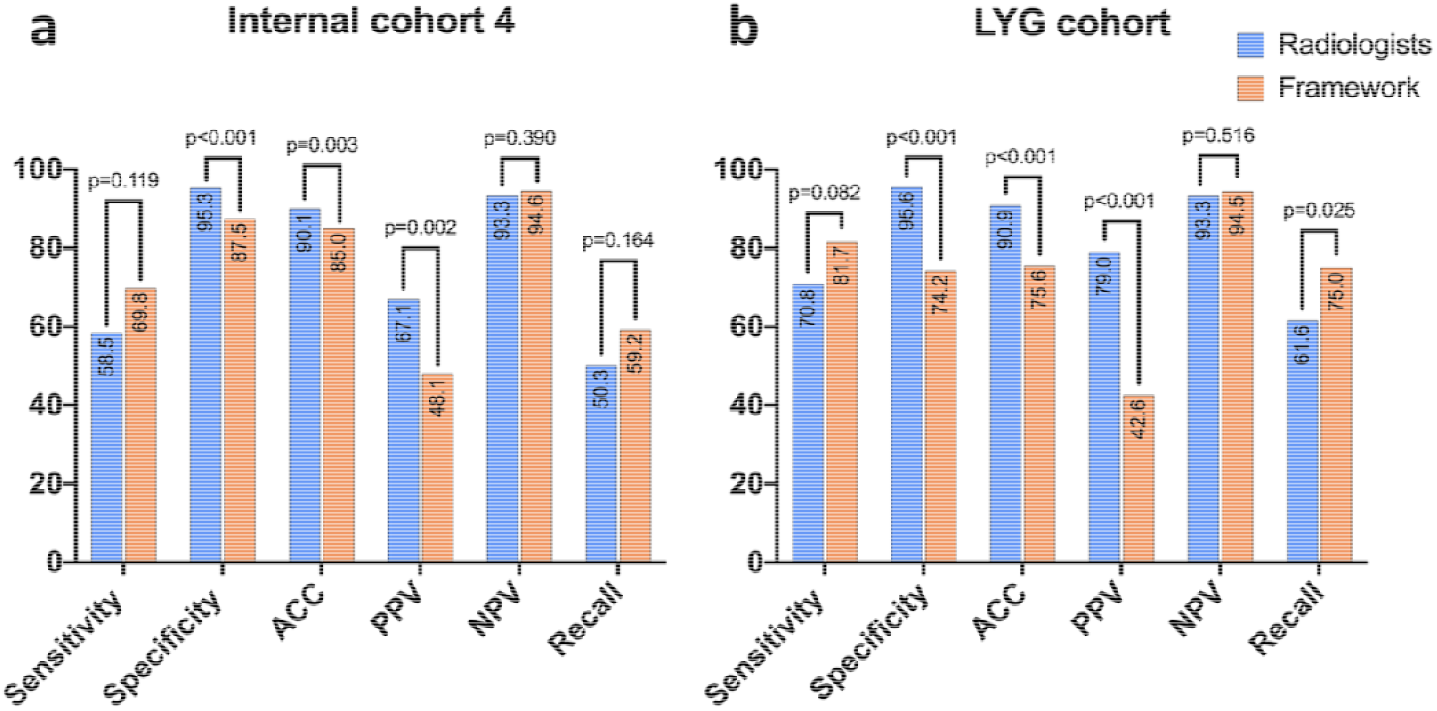
Comparison of the performances of the framework and the radiologists in Internal cohort 4 and LYG cohort. **a**, Performance in Internal cohort 4. The framework had improvement in sensitivity and NPV to that of the radiologists. **b**, Performance in external LYG cohort. Similar results can be found that the framework had likely higher sensitivity and NPV to those of the radiologists, and the specificity, ACC and PPV were significantly lower for the framework (p<0.05). ACC, accuracy; NPV, negative predictive value; PPV, positive predictive value.

A completely independent internal validation dataset (**Internal cohort 2**) was applied to test the framework, which contained 245 cases performing both head CTA and DSA during Apr. 1, 2017 and Dec. 31, 2017 in Jinling Hospital (**Table 1 and Extended Data Table 1**). The model reached accuracy, sensitivity, and specificity of 86.1%, 88.3%, and 84.3%, with a recall rate of 79.7% and FPs of 0.26/case (**Table 2**). Totally 30 aneurysms from 27 patients were missed, including 18 IAs in 15 patients with multiple IAs. The framework had high recall rate for aneurysms located in PCoA (93.9%), ACoA (96.6%) and ACA (90.9%), while lower recall rate in MCA (80.0%), PCA (66.7%), ICA (55.9%) and CA (50.0%); the most frequently missed aneurysms were tiny aneurysms (≤ 3 mm, recall rate of 43.8%), while it had high recall rate of 89.6% for those ≥ 3mm, 90.0% for those ≥ 5 mm, 100% for those ≥ 10 mm.

**Table 2.** Performance of the model in all cohorts

| **Cohort** | **Accuracy** | **Sensitivity** | **Specificity** | **PPV** | **NPV** | **Recall** | **FP** | **Dice** |
| --- | --- | --- | --- | --- | --- | --- | --- | --- |
| **Internal cohort 1** | 86.0%<br>(79.5%-90.7%) | 97.3%<br>(90.8%-99.3%) | 74.7%<br>(63.8%-83.1%) | 79.4%<br>(70.0%-86.4%) | 96.6%<br>(88.3%-99.1%) | 95.6%<br>(89.1%-98.3%) | 0.29<br>(0.23-0.37) | 0.752<br>(0.708-0.796) |
| **Internal cohort 2** | 86.1%<br>(81.2%-89.9%) | 88.3%<br>(81.0%-93.0%) | 84.3%<br>(77.2%-89.5%) | 82.4%<br>(74.5%-88.2%) | 89.7%<br>(83.2%-93.9%) | 79.7%<br>(72.5%-85.4%) | 0.26<br>(0.21-0.32) | 0.626<br>(0.568-0.684) |
| **Internal cohort 3** | 86.8%<br>(80.4%-91.3%) | 82.6%<br>(69.3%-90.9%) | 88.6%<br>(81.1%-93.3%) | 76.0%<br>(62.6%-85.7%) | 92.1%<br>(85.1%-95.9%) | 74.6%<br>(62.2%-83.9%) | 0.17<br>(0.12-0.24) | 0.587<br>(0.488-0.686) |
| **Internal cohort 4** | 85.0%<br>(81.1%-88.3%) | 69.8%<br>(56.5%-80.5%) | 87.5%<br>(83.5%-90.7%) | 48.1%<br>(37.3%-59.0%) | 94.6%<br>(91.4%-96.7%) | 59.2%<br>(47.5%-69.8%) | 0.20<br>(0.16-0.24) | 0.435<br>(0.339-0.531) |
| **Internal cohort 5** | 86.4%<br>(81.2%-90.4%) | 40.0%<br>(16.8%-68.7%) | 88.7%<br>(83.7%-92.4%) | 14.8%<br>(5.9%-32.5%) | 96.8%<br>(93.2%-98.5%) | 33.3%<br>(13.8%-60.9%) | 0.14<br>(0.10-0.20) | 0.298<br>(0.016-0.579) |
| **NBH cohort** | 80.1%<br>(74.2%-84.9%) | 82.1%<br>(67.3%-91.0%) | 79.7%<br>(73.0%-85.0%) | 47.8%<br>(36.3%-59.5%) | 95.1%<br>(90.3%-97.6%) | 72.3%<br>(58.2%-83.1%) | 0.27<br>(0.22-0.34) | 0.510<br>(0.409-0.611) |
| **TJ cohort** | 61.0%<br>(48.3%-72.4%) | 64.1%<br>(48.4%-77.3%) | 55.0%<br>(34.2%-74.2%) | 73.5%<br>(56.9%-85.4%) | 44.0%<br>(26.7%-62.9%) | 64.0%<br>(50.1%-75.9%) | 0.69<br>(0.57-0.80) | 0.404<br>(0.285-0.522) |
| **LYG cohort** | 75.6%<br>(70.6%-80.0%) | 81.7%<br>(70.1%-89.4%) | 74.2%<br>(68.5%-79.2%) | 42.6%<br>(34.0%-51.7%) | 94.5%<br>(90.5%-96.9%) | 75.0%<br>(64.2%-83.3%) | 0.32<br>(0.27-0.38) | 0.550<br>(0.465-0.634) |

*The data in parentheses are 95% confidence interval;*
*FP, false positive; NPV, negative predictive value; PPV, positive predictive value.*

Another completely independent external validation dataset from Nanjing Brain Hospital (**NBH cohort**) between Jan. 1, 2019 and Jul. 13, 2019 was collected to test the model’s generalizability, which contained 211 cases with both head CTA and DSA and 39 patients had IAs. The framework reached accuracy, sensitivity, and specificity of 80.1%, 82.1%, and 79.7%, with a recall rate of 72.3% and FPs of 0.27 (**Table 1**). Totally 13 aneurysms from 13 patients (including 6 IAs in 6 patients with multiple IAs) were missed. The framework had 100% recall rate for aneurysms located in ACoA, ACA, and PCA, while 63.3%, 80.0%, 66.7% and 62.5% for those in ICA, PCoA, MCA, and VBA. Similarly, it had a lower recall rate for tiny aneurysms (33.3%), while 82.9% recall rate for those ≥ 3, 90.5% for those ≥ 5mm, 100% for those ≥ 10 mm. There was no significant difference between the performance (sensitivity, specificity, and recall rate) in the internal and external validation cohorts (Internal cohort 2 and NBH cohort, *p*=0.394, 0.365 and 0.369, respectively).

### Comprehensive Analysis of the Performance of the Framework

In the process of model development, we found 31 occult cases that were CTA-negative but DSA-positive aneurysms, especially for tiny aneurysms or those located in the ophthalmic artery segments, which had been reported by some researchers^31^. These cases were abandoned in the **Internal cohort 1** while remaining in the validation set in our original designation for that they were hard to annotate in CTA source images. We wondered whether these cases could be detected by the framework given the magic power of DL. There are 39 aneurysms totally regarded as occult aneurysms (mean size: 2.0 (2.0, 2.5) mm, range: 1.0-4.0 mm). Most of these aneurysms were located in ICA (53.8%, 21/39), especially in the ophthalmic artery segments (57.1%, 12/21). Our framework detected 7 occult aneurysms with the mean size of 1.9 mm from 6 patients, among which 3 were located in ACoA, 1 in ACA, CA, ICA, and MCA, respectively.

Image quality affects the diagnostic performance of CTA, especially for small aneurysms. Herein we analyzed the level of tolerance of our framework to the image quality of head CTA. CTA image quality was rated on a four-point scale^32^, which is based on the degree of noise, vessel sharpness, and overall quality **(Extended Data Fig. 3)**. Further analysis was performed on another internal validation dataset (**Internal cohort 3**) including 151 patients. 47 patients had 60 aneurysms, while 104 patients had no aneurysms. There were 10, 43, 65, and 33 cases for the image quality scores 1-4, respectively. The results demonstrated that the sensitivities were 66.7%, 100%, 73.9% and 83.3% in the groups of score 1-4, and the corresponding specificities were 85.7%, 89.7%, 85.7%, and 92.6%, respectively **(Extended Data Fig. 4a\c)**. There was no significant difference among the four groups (all *p*>0.05), which meant the model had a high tolerance to image quality. However, another truth is that with the advance of CT techniques and optimized CTA protocols, poor quality images (score 1) are rare. We can barely collect 10 cases among all head CTA cases, which means the model is qualified for further application.

**Fig.3.**
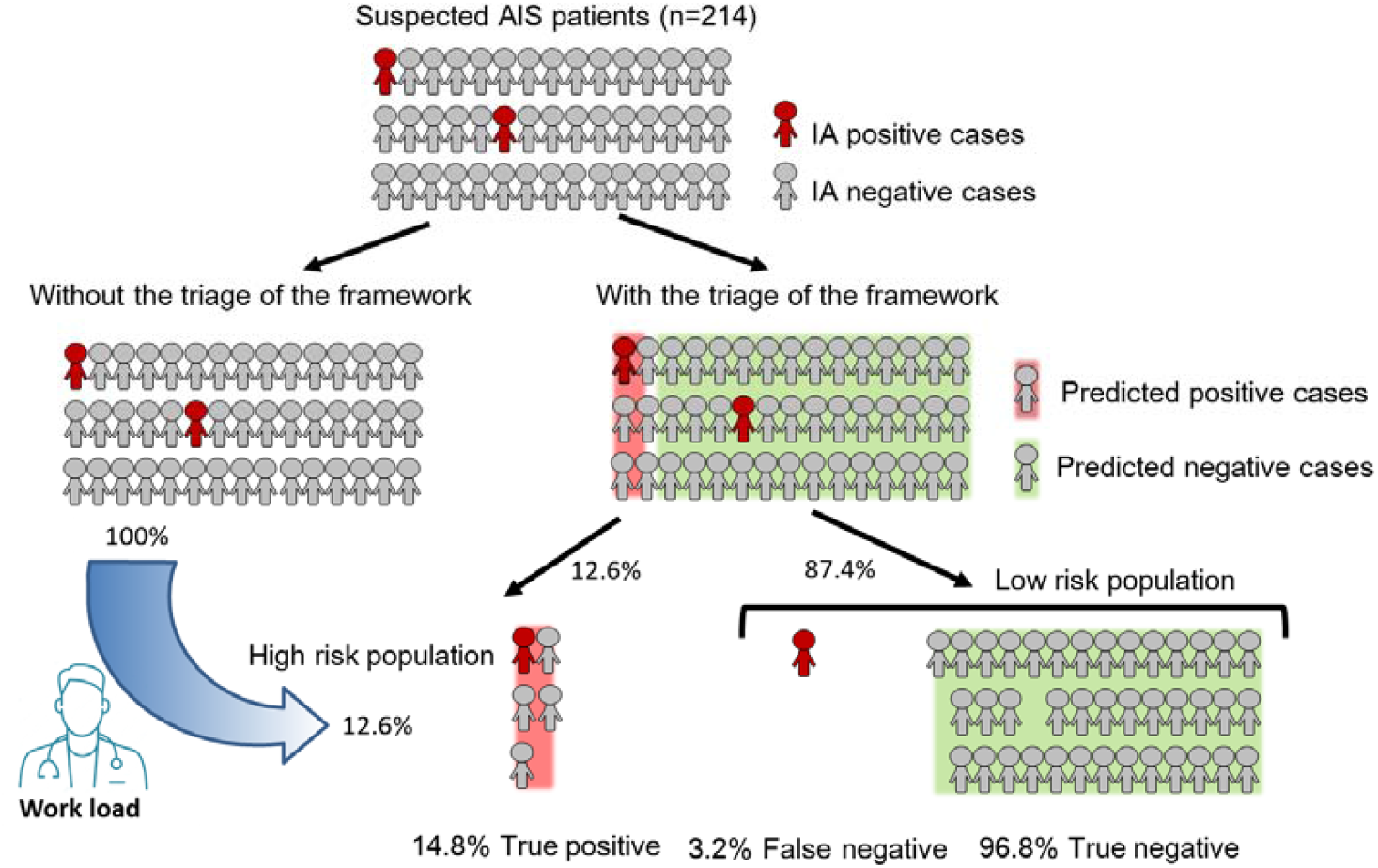
Impact of the proposed framework on clinical practice for patients with suspicious acute ischemic stroke in emergent department. In the cohort of patients with suspected AIS (**Internal cohort 5**), who were prescribed to undergo head CTA examination, 87.4% patients diagnosed as aneurysm-negative cases by our framework, among which 96.8% of the patients were true-negative patients, demonstrating excellent confidence in identifying negative cases by our framework. As a result, only 12.6% of patients were categorized as high-risk group, to whom the radiologists can pay intense attention and reduce their workload in detecting aneurysm for AIS patients. AIS, acute ischemic stroke; IA, intracranial aneurysm.

Manufacturers can be a factor to affect the performance of the framework, so the Tianjin dataset (**TJ cohort**) between Jan. 1, 2013 and Dec. 31, 2018 was employed, which had both head CTA and DSA from 3 different manufacturers to elucidate the issue. CTA was acquired by GE Revolution in 13 patents (10 patients with IAs), by Siemens SOMATOM Definition Flash in 21 (18 patients with IAs), by Toshiba Aquilion ONE in 25 (11 patients with IAs). The results demonstrated that the sensitivity and specificity were 70.0%, 66.7%; 72.2%, 66.7%; 45.5%, 50% for GE, SIEMENS and Toshiba, respectively, while without significant differences (all *p*>0.05) **(Extended Data Fig. 4b/c)**.

### Radiology Expert Analysis of the Error Modes

For knowing the underlying causes of misclassified cases in our developed framework, neuroradiologists analyzed the cases in the **Internal cohorts 2, 3, NBH cohort** and **TJ cohort**. The misdiagnosed cases (13 in **Internal cohort 2**, 8 in **Internal cohort 3**, 7 in **NBH cohort** and 14 in **TJ cohort**) were found between the framework diagnosis and the DSA findings (**Extended Data Table 2a**). Since that misdiagnosis would lead to serious consequences, we only focus on these cases rather than the FPs. Several reasons can attribute to misdiagnosed cases, including (1) Tiny aneurysms (≤ 3 mm); (2) Uncommon shape of aneurysms; (3) Aneurysms located in uncommon locations (such as the posterior inferior cerebellar artery); (4) Inappropriate contrast agent protocols that result in poor artery enhancement or marked intracranial vein enhancement; (5) Other unexplained reasons (**Extended Data Table 2b)**. We defined unexplained reasons for that these aneurysms were obvious for radiologists to catch, while were missed by our framework, which was mainly found in the **TJ cohort** and **NBH cohort** and may be contributed to different manufacturers or CTA scan protocols. And another case with unexplained reason was seen in **Internal cohort 1** and the reason might be the overshooting bone subtraction (a representative missed case is presented in **Extended Data Fig. 5)**.

### Clinical Application in Routine Practice and Comparison with Radiologists

In order to clearly understand the performances of the framework against radiologists in clinical setting, we designed an experiment to compare the performance of the framework to those of 6 board-certified radiologists (2 resident radiologists, F.X. and Y.X.; 2 attending radiologists, L.W. and X.L.Z.; 2 assistant director radiologists, J.L. and Y.E.Z.) in reading consecutive real-world cases undergoing head or head/neck CTA, in one internal dataset from Jun. 1, 2019 and Jul. 31, 2019 in Jinling Hospital (**Internal cohort 4**) and one external dataset from Aug. 1, 2018 to Sep. 31, 2019 in Lianyungang First People’s Hospital (**LYG cohort**). The prevalence of IAs is distinct in general and SAH populations^1,2^, which acts as an important clue for detecting IAs for radiologists. Therefore, we analyzed the performances of the framework and radiologists in the whole group, SAH subgroup, and non-SAH subgroup, respectively.

For **Internal cohort 4**, the microaverage sensitivity and specificity were 58.5%, 95.3%; 66.7%, 95.4%; 56.6%, 95.3% for the radiologists in the whole group, SAH group and non-SAH group, respectively. The radiologists seemed to had higher positive predictive value (PPV) in the SAH group than the non-SAH group (88.9% vs 62.9%, *p*=0.358), and vice versa for negative predictive value (NPV) (83.7% vs 93.9%, *p*=0.285), which demonstrated the additional value of SAH for diagnosis of IAs^9^. The recall rates were 50.3%, 54.8% and 49.1% for radiologists in the three groups. For the framework, it had likely higher sensitivity (69.8% (*p*=0.224), 80.0% (*p*>0.999) and 67.4% (*p*=0.268)) and NPV (94.6% (*p*=490), 88.9% (*p*>0.999), and 95.0% (*p*=0.545)) (**Fig. 2 and Extended Data Table 3)**. And the framework had a comparative recall rate of 59.2% (*p*=0.312), 64.3% (*p*=0.699) and 57.9% (*p*=0.348) **(Extended Data Table 3)**. In the general, our framework had likely higher sensitivity and NVP but without significant differences in the whole group, SAH and non-SAH subgroups, while the specificities were significantly lower than human radiologists (*p*<0.001). Compared to the radiologists, the framework showed non-inferiority at a pre-specified 5% margin for sensitivity in the whole group and non-SAH subgroup. The mean diagnosis time per examination microaveraged across clinicians was 30.1 seconds (95% CI: 29.2-31.0 seconds). While the framework took 18.2 seconds (95% CI: 17.9-18.4 seconds) per case and was significantly faster than the radiologists (*p*<0.001).

Similar results can be found in **LYG cohort**, the microaverage sensitivity and specificity were 70.8%, 95.6%; 81.3%, 96.2%; 63.3%, 95.5% for radiologists in the whole group, SAH subgroup and non-SAH subgroup, respectively. PPV was higher in the SAH group (96.1% vs 67.9%, *p*<0.001) than non-SAH group, and vice versa for NPV (81.9% vs 94.6%, *p*=0.026). The recall rates were 61.6%, 72.5% and 53.9%, respectively (**Fig. 2 and Extended Data Table 3**). The framework had likely higher sensitivity (81.7% (*p*=0.082), 92.0% (*p*=0.306) and 74.3% (*p*=0.209)), NPV (94.5% (*p*=0.516), 88.9% (*p*=0.683), and 95.1% (*p*=0.772)) and recall rate (75.0% (*p*=0.025), 84.8% (*p*=0.131) and 67.4% (*p*=0.095)). Compared to radiologists, the framework demonstrated an improvement in sensitivity with noninferiority at a pre-specified 5% margin. The microaverage diagnosis time was 27.1seconds (95% CI: 26.3-28.0 secondes). And the framework took 19.6 seconds (95% CI: 19.3-20.0 seconds) per examination with a significant difference (*p*=0.001).

### Clinical Application in the Workup of AIS in Emergent Department

The results above demonstrated that our framework had likely higher recall rate, sensitivity, and NPV than the radiologists, which may have the potential for complementary implication in clinical practice. Inspired by this finding, we wonder whether this framework can work well when excluding the control cases to reduce workload in a specific clinical setting. The newly published guideline has strongly recommended CTA (with CT Perfusion) for AIS patients (Class I and level A) for selecting candidates for mechanical thrombectomy^14,15^. Radiologists are expected to detect stenotic and occlusive lesions and, as a result, often overlook the aneurysms. Administration of antiplatelet or anticoagulant therapy is often recommended, which exposed the patients to the risk of aneurysm rupture if they have IAs^13^. Therefore, radiologists should also pay special attention to the presence of IAs in this setting. Our framework is potential to apply in this population to exclude patients without IAs with high confidence so that high risk patients with IAs could be focused more intensively. Another 332 consecutive patients from the emergency department of Jinling Hospital underwent emergent head CTA examination from Jul. 1, 2018 to Jul. 31, 2019 were enrolled. The inclusion criteria are the patients with suspected AIS referred to CTA. The exclusion criteria were head trauma or surgery history (n=29), poor image quality (n=6) and intracranial hemorrhage (n=83), among which 41 patients had SAH. Finally, 214 patients suspicious AIS were included (10 patients with IAs) (**Internal cohort 5**). The results demonstrated that our framework had a specificity of 88.7% while the sensitivity was relatively low (40.0%), and the NPV was 96.8%. From this result, we can assume that, with the triage of the framework, 87.4% of patients were predicted as negative, among which 96.8% predicted-negative cases are true-negatives, and the other 12.6% were predicted as high-risk group with the aneurysm. Therefore, radiologists can focus on these patients with more intense attention in order to improve workflow and reduce workload (**Fig. 3**).

On the other hand, the 3.2% FN patients cannot be ignored. In this experiment, 8 aneurysms from 7 patients were missed, among which all 7 IAs in 6 patients were misdiagnosed. All missed aneurysms were small and mainly located in ICA (5/8, 62.5%) and MCA (2/8, 25.0%).

## Discussion

In this study, we proposed and developed a novel DL framework to automatically predict and segment IAs in bone-removal CTA images and conducted a comprehensive analysis of the influence of occult cases, image quality and manufacturers, which indicated the framework’s high tolerance. The validation process in the simulated real-world scenarios proved that the framework had higher sensitivity and NPV than radiologists. Inspired by this, we further validated the framework in the suspected AIS clinical scenario, which demonstrated that the framework could exclude IA-negative cases with high confidence and help prioritize the clinical workflow to reduce the workload and thus the radiologists could focus on the high-risk patients. We believe our proposed framework is innovatively designed and the study had a complete workflow of development and validation procedures from laboratory to real-world settings for clinical application. While implementation studies are warranted to develop appropriate and effective radiologists’ alerts for the potentially critical finding of IAs and to assess their impact on reducing time to treatment.

There are several reasons contributing to the complexity of the IAs detection task. Firstly, arterial visualization of CTA images is easily affected by the enhanced veins. Secondly, the prevalence of IAs in the general population and the SAH population is significantly different (2-3% vs. 80-85%)^33,34^. Thirdly, the IA detection procedure includes two steps for radiologists, the first step is to identify cerebral arteries and then to recognize abnormal dilation (aneurysm), which is different from diagnosing solid lesions such as lung cancer^35^, retinal diseases^19^, and thyroid cancer^20^. The peculiarity of head CTA has resulted in little efforts that apply supervised learning to IA classification and detection. Because of the reasons, CAD of CTA to detect IAs has been rarely reported, and only two studies can be found^25,26^. The lack of a reference standard, external validation data or focusing only on non-ruptured aneurysms ≥ 3 mm limit the generalization and further application of their models in the clinical setting. In order to solve these issues, we enrolled a large number (n=1177) of CTA cases for training the novelly designed DL framework for high robust, which had been verified by DSA, the gold standard for IAs diagnosis, especially for small ones. Besides, we collected external datasets from other hospitals for validation of generalizability. Other factors that influenced the model performance were also taken into consideration, including image quality, occult IAs, and different manufacturers, and some of the cohorts had also been verified by DSA.

Our study also highlights the potential clinical application of the framework in a suspected AIS cohort, for which head CTA is recommended and the result demonstrated that the framework could reduce radiologists’ workload by triage. With all these efforts, our work provided a relatively complete and innovative insight into the development and validation of the framework for automatic prediction and segmentation of IAs.

Several approaches adopted to improve framework performance. First, the proposed model was built based on the U-Net^30^, a well-proven network structure that has been widely used in medical image segmentation. Second, we used the Basic Block instead of the stacked convolution layer, thus the performance of the deep conventional network was boosted by the residual connection. Last, the dual attention block^30^ was employed to enforce the network to focus on the informative region and features. We also compared the performances of the framework to that of the most frequently employed 3D U-Net^36^ using the same training and testing data (**Internal cohort 1**), and our framework had significantly higher performance (**Extended Data Table 4)**.

Another strength of our proposed framework is that all CTA studies in the training, some of the internal and external validation cohorts were demonstrated by DSA results, which guaranteed our model’s performance. Although inter-grader variability is a well-known issue in many settings including IAs diagnosis in CTA^37-39^, human interpretation is still used as the reference standard in some studies^24-26^. Unambiguous interpretations which are called noisy labels can lead to an obviously biased performance of the model^40,41^. Therefore, it is necessary and urgent to use DSA, gold standard for IAs detection in machine learning based CAD studies.

We also collected consecutive CTA images from other medical centers to test the generalizability of this AI tool. Consequently, our framework is potential to overcome some difficulties in the clinical practice of aneurysm detection, including a large number of diagnostic errors, an enormous waste of resources, and inefficiencies in the workflow^42,43^. Given these model results, readers had the option to take it into consideration or disregard it based on their own judgment. With this approach to medical imaging, the framework offers the visualized results of suspected IAs on CTA images at slice-level, in a fashion similar to clinical practice. As a result, an interpretable representation is particularly useful in difficult and ambiguous cases. Such cases are common in clinics and even specialized neuroradiology practitioners are difficult to detect.

Even though we have proved the high performance of the framework, and the framework can be run in real-time within or across entire hospital systems, we have to note that the adverse effects on radiologists’ workload and behaviors. For example, radiologists must respond to the FP predictions, which may result in increasing fatigue of radiologists and blunt human responsiveness to TP predictions by the framework. In contrast, we have to avoid the unfavorable result of clinician overreliance on automated systems. On the other hand, there were still some visible aneurysms that were ignored by the framework, such as those located in ICA and MCA, and tiny aneurysms, for which the radiologists must pay attention when augmenting with the tool.

The limitations mainly lay in the relatively small sample of positive cases in the validation cohorts, which cannot be avoided because of the relatively lower prevalence of IAs. The second limitation is the data curation process, where AVM/AVF and head trauma data were abandoned. This means the algorithm may not perform as well for images with subtle findings that a majority of radiologists would not identify. Thirdly, the performance of radiologists augmented by the framework is not discussed in this study for that a long journey of permission of device installation exists. However, we can assume that the human augmented performance would be better than that of without augmentation, which has been shown in one study by Park et al^25^. Finally, we have not conducted a prospective multicenter controlled experiment to validate the framework in clinical scenarios.

Notwithstanding these concerns, our proposed novel DL based framework for automated detection of IAs had higher sensitivity and recall rate compared with radiologists and could reduce radiologists’ workload. Several aspects of clinical effectiveness should be measured and tracked, including patient outcomes and costs. Therefore, further worthwhile consideration is how to best integrate the model with the routine radiology interpreting workflow, how to best leverage the complementary strengths of the DL framework and clinician gestalt and experience.

## Online methods

### Ethics and information governance

This retrospective study was approved by the Institutional Review Board of Jinling Hospital, Medical School of Nanjing University, with a waiver of written informed consent. Only de-identified retrospective data were used for research, without the active involvement of patients.

### Data

We did a retrospective, multicohort, diagnostic study using head or head/neck bone-removal CTA image sets from four hospitals in China (Jinling Hospital, Tianjin First Central Hospital, Lianyungang First People’s Hospital, and Nanjing Brain Hospital). As listed in **Table 1** and **Extended Data Table 1**, 8 cohorts were applied to devise and validate the framework.

**Internal cohort 1** encompassed patients who underwent CTA examinations and afterward verified by cerebral digital subtraction angiography (DSA) within 30 days in Jinling Hospital, Nanjing, China between Jun. 1, 2009 and Mar. 31, 2017. The data were then shuffled and separated into a training/tunning/test sets. The tunning set consisted of 100 cases (50 cases with aneurysms and 50 non-aneurysm controls), and the testing set had 150 cases with 50% cases having IAs. The tunning set was used to evaluate model performance at the end of each epoch during training and for hyper-parameter optimization, and the test set was a held-out set of images used for evaluation of the trained models, never used by the algorithm during training or validation.

For internal validation sets, consecutive patients undergoing CTA examinations verified by DSA from Apr. 1, 2017 to Dec. 31, 2017 in Jinling Hospital (**Internal cohort 2**) were used for internal validation. Consecutive patients undergoing CTA examinations verified by DSA during 2018 (**Internal cohort 3**) were collected for validation of the effect of image quality. Patients who underwent head or head/neck CTA while without DSA restraints from Jun. 1, 2019 to Jul. 31, 2019 in Jinling Hospital (**Internal cohort 4**) were obtained for simulated real-world validation and human-model comparison. Patients who suspected of AIS from Jul. 1 2018 to Jul. 31 2019 (**Internal cohort 5**) were collected for the function validation whether the confident screen of aneurysm-negative cases can reduce radiologists’ workload.

For external validation, DSA-verified consecutive eligible CTA cases from Jan. 1 2019 to Jul. 31 2019 in Nanjing Brain Hospital (**NBH cohort**) were enrolled for the effect validation of external data. DSA-verified consecutive eligible CTA cases from Tianjin First Central Hospital were collected for validation of the effect of different manufacturers in 2013-2018 (**TJ cohort**) to the framework, which contained 3 different manufacturers including GE Revolution, Siemens SOMATOM Definition Flash and Toshiba Aquilion ONE. Consecutive eligible CTA cases from Aug. 1 2018 to Sep. 31 2019 in Lianyungang First People’s Hospital (**LYG cohort**) were collected for simulated real-world validation and human-model comparison (**Extended Data Table 1**).

All CTA images extracted from the 4 hospitals were in DICOM format. Four multidetector CT scanners (SOMATOM Definition and SOMATOM Definition Flash, GE Revolution CT, Toshiba Aquilion ONE) were used to generate source CTA images and all images were processed in a workstation (Syngo 2008G; Siemens) with the bone voxels removed by software (Neuro DSA application) in core lab. The bone-removed DICOM images were used to annotate and training for models.

For patients who underwent CTA and verified by DSA, the interval was less than 30 days. A dedicated curation process was only conducted in the training dataset (**Internal cohort 1**) for high-quality images in the training set, and the exclusion criteria are as following: a). patients who underwent DSA before head CTA; b). patients with more than 30 days interval between CTA and DSA; c). patients who had surgical clips, coils, catheters, or other surgical hardware; d). patients with arteriovenous malformation/fistula (AVM/AVF), Moyamoya disease, arterial occlusive diseases and other vasculopathies that affect the structure of intracranial vasculature; e). patients with incomplete image data, images of poor quality and unavailable images; f). patients with IA in DSA but undetectable in CTA images. The control group included those underwent CTA and DSA and all images were reviewed and were free of abnormal cerebral vasculopathies. Patients with infundibular dilations and vascular stricture lesions were also included as controls considering the negligible influence. **Extended Data Fig. 1** shows the flowchart of this study.

While in the validation cohorts with DSA verification, we only excluded patients who met exclusion criteria a), b), c), e), and those with AVM/AVF (This vasculopathy was obvious to detect while had a morphological influence on the intracranial arteries.) to test the applicability of the proposed framework in a real-world clinical scenario. In those without DSA verification, we excluded patients who met exclusion criteria: c), e), and those with AVM/AVF.

### Radiologist annotations

The presence and locations of IAs in each patient was determined by DSA (**the gold standard**), if available. Specifically, the spatial resolution of cerebral CTA was inferior to that of DSA, therefore the locations of IA were reviewed by three specialized neuroradiologists (Z.S., S.L. and C.S.Z. with 3, 8 and 13 years experiences in neuroradiology interpretation, respectively) with reference to the DSA images to establish the final standard.

For the patients without DSA verification, two specialized neuroradiologists (C.S.Z. and S.L.) had access to all of the Digital Imaging and Communications in Medicine (DICOM) series, original reports, and clinical histories, as well as previous and follow-up examinations during interpretation to establish the most possible reference standard labels (**the silver standard**) and they had rights to exclude CTA of poor quality and incomplete images that were not satisfied for diagnosis as well as CTA cases that were of post-procedure. In the case of disagreement between the two observers, consensus was reached in a joint reading with the assist of a senior neuroradiologist (L.J.Z. with 19 years of neuroimaging experience) and then the majority vote of 3 radiologists established reference standard labels.

The explicit locations of IA for all examinations in the training set were determined by DSA, and one annotationist (Z.S.) matched the location into the corresponding site on bone-removal CTA of DICOM series in pixel-wise with Mimics software (Version 16.0). The annotationist had access to all of the DICOM series and the final standard to identify the accurate location of IA on CTA. The identified aneurysms were manually segmented using MIMICS (Version 16.0) on slices that contain IAs in bone-removal CTA.

### Model development

In this study, we developed a 3-dimensional (3-D) CNN called DAResUNet for the segmentation of IAs from bone-removal CTA images. DAResUNet is a CNN with an encoder-decoder architecture similar to 3D U-Net^30^, which contains an encoding module (encoder) for abstracting contextual information and symmetric decoding module (decoder) for expanding the encoded features to a full-resolution map with the same size and dimensionality as the input volume. We adopted residual blocks^28^ to replace the original convolution blocks of U-Net to ensure stable training when the depth of the network is significantly increased. Besides, dilated convolutions were used in the top layer of the encoder to enlarge the receptive field of the network, as we only performed down-sampling 3 times. To enhance the performance of the network by exploring long-range contextual information, we embed a dual attention^29^ module between the encoder and the decoder.

The size of the input volume of the DAResUNet was 80×80×80, which is large enough to enclose the majority of IAs. 3D image patches with the above size were randomly sampled from the entire CTA volume during training. To balance the number of training samples containing and not containing aneurysms, sampled patches have a 50% probability to contain an aneurysm. Before patch sampling, data augmentations such as rotation, scaling and flipping were applied to CTA scans.

Before reaching the network, inputs were clipped to [0, 900] Hounsfield units and then normalized to [-1, 1]. The network was trained to optimize a weighted sum of a binary cross-entropy loss and a Dice loss. The Adam optimizer was used by setting the momentum and weight decay coefficients to 0.9 and 0.0001, respectively. We employed a poly learning rate policy, where the initial learning rate is multiplied by 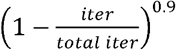 after each iteration. The initial learning rate was 0.0001 and the number of training epo1hs was 100. During each training epoch, about 50 patches were sampled from each CTA scan.

In the inference stage (**Fig. 1b**), the segmentation prediction of the whole volume was generated by merging the prediction of uniformly sampled patches. Two adjacent patches may have 1/8 overlap, in other words, the strides along the three axes were all 40. For each voxel, we used the highest probability from all enclosing patches as its final prediction.

### Image quality dependence of the framework performance

In this study, we randomly collected 152 consecutive CTA cases acquired in 2018 in Jinling Hospital, all of which had been verified by DSA and overviewed by 3 specialized neuroradiologists (Z.S., S.L., and C.S.Z.) to determine the diagnosis of IAs. Image quality was rated using multiplanar reconstructions, maximum intensity projections, and volume-rendered reformatted images^27^. Qualitative image scoring was performed independently by two neuroradiologists (Z.S. and S.L.). In case of disagreement between both readers, consensus was reached in a joint reading to determine the final image quality score.

### Human-model comparison experiment

It is relatively easier to develop a DL framework in a curated experimental environment than applying it in an ethical, legal and morally responsible manner within a real-world healthcare setting. Therefore, we designed a validation process that simulates the real-world clinical procedures by applying the framework in **Internal cohort 4** and **LYG cohort** which highlights the true context of the full breadth of CT scanning presented to clinicians from real-life clinical practice.

We performed a diagnostic accuracy study comparing performance metrics of radiologists with different years of experience and the model. Six board-certified radiologists (2 resident radiologists, F.X. and Y.X. with 4 years of working experience; 2 attending radiologists, L.W. and X.L.Z. with 7 years of working experience; and 2 assistant director radiologists, J.L. and Y.E.Z. with 11 and 13 years of working experience) participating in the study were asked to interpret the CTA images and identify the presence of IAs. None of the 6 radiologists was involved in the procedure of determining the location of IA as the reference standard. The 6 radiologists read all the CTA images from **Internal cohort 4** and **LYG cohort**. Acquired CTA image series were manually transferred to a dedicated workstation for review (Multi-Modality Workplace; Siemens Healthcare). CTA images were generated with a workstation (Syngo 2008G; Siemens) with the bone voxels removed by software (Neuro DSA application). The radiologists were blinded to clinical data as well as reference standard labels and independently analyzed all cerebral CTA images by using source images, maximum intensity projections, multiplanar reformations, volume-rendering reformatted images and target vessel reformation to determine the presence and locations of aneurysms, a way that simulates the real procedures of clinical practice. They were allowed to adjust, rotate, and reformat 3D images at the workstation to optimally view the presence and location of an individual aneurysm. While the model only used the bone-removal source images to generate IA predictions.

They read independently in a diagnostic reading room, all using the same high-definition monitor (1920 × 1200 pixels) displaying CTA examinations on the dedicated workstation (Multi-Modality Workplace; Siemens Healthcare). The microaverage of sensitivity, specificity, accuracy, PPV and NPV across all radiologists were computed by measuring each statistic pertaining to the total number of true-positive, true-negative, false-positive and false-negative results.

### Statistical analysis

Quantitative variables were expressed as mean ± SD if normally distributed, while median and inter-quartile range (IQR) was used when non-normally distributed data. Categorical variables were expressed as frequencies or percentages. The segmentation framework detects the potential aneurysm lesions in the head/head-neck CTA image data. The accuracy, sensitivity, specificity, PPV, and NPV of the correct display of patients with IAs by the framework were evaluated separately in each cohort and the 95% Wilson score confidence intervals were used to assess the variability in the estimates for sensitivity and specificity. The performances of radiologists were calculated in **Internal cohort 4** and **LYG cohort**. We also calculated likelihood ratios for positive and negative results. To assess model performance against that those of 6 radiologists, we used a 2-sided Pearson’s chi-squared test to evaluate whether there were significant differences in specificity, sensitivity, accuracy, PPV, and NPV between the framework and radiologists. For comparisons with radiologists, the choice of superiority or non-inferiority was based on what seemed attainable from simulations conducted in **Internal cohort 4** and **LYG cohort**. The confidence limits of the difference were based on Gart and Nam’s score method with skewness correction. For non-inferiority comparisons, a 5% absolute margin was pre-specified before the test set was inspected. Statistical analyses were conducted with SPSS Statistics (version 22.0.0, IBM SPSS Statistics, Armonk, New York), R (version 3.5.2, R Foundation for Statistical Computing, Vienna, Austria) and NCSS (Version 12, LLC, Kaysville, Utah,). We used a statistical significance threshold of 0.05.

## Data Availability

All the data are available from the authors upon reasonable request.

## Data availability

All the data are available from the authors upon reasonable request.

## Acknowledgements

This work was partly supported by the National Key Research and Development Program of China (2017YFC0113400 for L.J.Z.) and Key Projects of the National Natural Science Foundation of China (81830057 for L.J.Z.).

We thank Drs. Fei Xia, Yuan Xie, Li Wang, Xiaolei Zhang, Jia Liu, and Yan’e Zhao for their effort in human interpretation of CTA examinations in Internal cohort 4 and LYG cohort; Xiang Kong, Wei Zhang, Li Qi, Weiwei Huang, Mengdi Li for DICOM data archive; Mengjie Lu for statistical analysis support and all those who had made contribution to the publication of the work.

## Author Contributions

L.J.Z., G.M.L., X.L.L. initiated the project and the collaboration. C.W.P., H.W., X.L.L., Y.Z.Y. developed the network architectures, training and testing setup. L.J.Z., Z.S., C.S.Z., X.L.L. designed the clinical setup. Z.S., C.S.Z., S.L. S.X., Y.G., Y.G.Z., C.C.M., X.C., B.H., W.D.X. created the dataset and defined clinical labels. C.W.P., H.W., contributed to the software engineering. Z.S., C.S.Z., S.L., S.X., Y.G., X.C. created the database. L.J.Z., C.S.Z., S.L., Z.S. contributed clinical expertise. Z.S., L.M., M.J.L., analyzed the data. L.J.Z., G.M.L., X.L.L. managed the project. Z.S., C.W.P., L.J.Z. wrote the paper. L.J.Z. contributed to the uncertainty estimation.

